# Frequent tumor burden monitoring of esophageal squamous cell carcinoma with circulating tumor DNA using individually designed digital PCR

**DOI:** 10.1101/2020.05.01.20087106

**Authors:** T. Iwaya, F. Endo, M. Yaegashi, N. Sasaki, R. Fujisawa, H. Hiraki, Y. Akiyama, A. Sasaki, Y. Suzuki, M. Masuda, T. Yamada, F. Takahashi, T. Tokino, Y. Sasaki, S.S. Nishizuka

## Abstract

**Background:** Circulating tumor DNA (ctDNA) test has not yet been an established tool for monitoring cancer. Sensitive, yet affordable methods should allow frequent ctDNA monitoring that can assist in clinical management.

**Patients and Methods:** This prospective observational study was conducted in a total of 36 patients with Stage I to IV esophageal squamous cell cancer (ESCC) were enrolled between September 1, 2015 and February 28, 2018. We investigated whether frequent ctDNA monitoring during treatment followed by routine surveillance by digital PCR (dPCR) using tumor-specific mutations offers clinical validity in daily practice for ESCC patients.

**Results:** Mutation screening of tumors from analyzable 35 patients using a specifically-designed "SCC panel" revealed 221 mutations with variant allele frequency (VAF) >2%. VAF of ctDNA was informative in 34 patients surveillance by dPCR using 58 mutations (1-3 per patient). A total of 569 plasma samples at 332 time points for ctDNA testing were evaluated. In pretreatment plasma, the average VAF was higher in advanced stages than earlier stages (*P* < .0001); tumor volume was also higher for higher VAF (*r* = 0.71). The ctDNA-positive rate in the pretreatment plasma of stage II or higher was 85.2% (23/27) whereas 85.7% (6/7) stage I were below the detection limit. Ninety-one *%* (10/11) patients whose ctDNA increased during chemotherapy showed disease progression. Among patients who recurred, ctDNA elevated with a median lead time of 149 days to the imaging diagnosis. Patients with decreased ctDNA within 3 months of initial treatment (n = 10) showed significantly better outcomes than did patients with ctDNA-positive (n = 11; *P* < .0001, HR 0.10, 95% CI, 0.03-0.30).

**Conclusions:** Our results indicate that frequent tumor burden monitoring using a small number of tumor-specific ctDNAs by dPCR enables prediction of relapse and chemotherapeutic efficacy, as well as relapse-free corroboration in management of ESCC patients.

## Introduction

Esophageal squamous cell carcinoma (ESCC), the most common esophageal cancer subtype in East Asia, has a poor prognosis and <20% 5-year overall survival rate [1]. At present, multidisciplinary treatments including surgery, chemotherapy, and radiotherapy are widely used to improve prognosis and symptoms for ESCC patients [2]. For treatment selection and recurrence detection, computed tomography (CT) scan and serum tumor markers have been standard approaches. However, accurate diagnosis is often difficult using these diagnostic modalities, particularly for frequent and accurate monitoring of tumor burden [3, 4]. Therefore, the development of new tumor markers is crucial to ensure that effective therapies are applied in a timely manner and that accurate diagnosis of relapse, evaluation of chemotherapeutic efficacy, and relapse-free corroboration can be determined.

Analysis of circulating tumor DNA (ctDNA) has demonstrated promising results for monitoring of several cancer types for relapse prediction and evaluation of treatment efficacy [5–10]. At present, few studies have demonstrated improved patient outcomes or economic advantages of ctDNA monitoring relative to monitoring methods currently used in daily practice [11, 12]. Although next generation sequencing (NGS) can detect mutations in multiple genes, this technique requires costly multiple sample/library preparation and analytical processes. Moreover, NGS is not designed in principle to target sequences that have low variant allele frequency (VAF). When the target VAF is <1%, as is seen for the majority of ctDNAs, intrinsic sequencing errors must be eliminated via techniques such as *in silico* error suppression [13]. Digital PCR (dPCR) was originally developed to quantitatively measure rare variants among large numbers of wild-type sequences [14]. Indeed, detection of VAF as low as 0.001% can be achieved by dPCR, suggesting that this technique can offer stable detection of rare variants when the VAF is above these levels [15]. Validated primer/probe sets allow numerous time-point assays to be performed for the same patient as well as for patients who share the same mutation. In addition, no substantial bioinformatic resources for dPCR analysis are needed to generate outputs given the simple nature of the PCR assay.

Here we demonstrate that frequent monitoring of ctDNA can offer timely treatment outcome information and provide potential opportunities for early intervention during the treatment course of ESCC patients.

## Materials and Methods

### Patients and sample collection

This study was approved by the Institutional Review Board of Iwate Medical University (IRB# HGH27-16) and was registered in the UMIN Clinical Trial Registry (UMIN000038724). Between September 24, 2015 and February 13, 2018, 36 histologically confirmed ESCC patients were enrolled (Stage I/II/III/IV: 8/3/20/5, respectively; supplementary Figure S1). Written informed consent was obtained from all patients. Patient characteristics are listed in supplementary Table S1. DNA was extracted from tumor tissues, peripheral blood mononuclear cells (PBMC) and a series of plasma samples. Methodology for DNA extraction from each type of sample is available in supplementary Methods and Figure S2.

### Panel sequencing

Tumor and corresponding PBMC DNA were subjected to amplicon sequencing using the Ion PGM™ system with a specifically designed SCC panel covering 617 exons of 31 genes that are frequently altered in ESCC and head and neck squamous cell carcinomas (supplementary Methods and Table S2). Sequencing analysis of the *TP53* gene in plasma DNA was also performed for patients having Stage IB or more advanced disease (supplementary Methods).

### Design and validation of dPCR probe and primer

The dPCR assay was used to detect rare variants in the primary tumor and quantitative monitoring of ctDNA (supplementary Methods and Figure S3) [16]. To investigate ctDNA by dPCR analysis, specific primers and probes labeled for wild type and mutant alleles were specifically designed for each mutation identified in primary tumors. From one to three mutations per patient with VAF higher than 10% in primary tumors were prioritized for dPCR analysis. Using Hypercool Primer & Probe™ technology (Nihon Gene Research Laboratories, Sendai, Japan), the appropriate size (<60-70 bp) of PCR products could be designed by using 5-methyl-dC and 2-amino-dA base modifications to adjust the *Tm* value. Prior to use in ctDNA monitoring, the synthetized primer and probe sets for each mutation were validated using corresponding primary tumor DNA.

### ctDNA dynamics in clinical time course

Longitudinal VAFs of ctDNA were plotted on a time course along with the imaging diagnosis, therapy types, and serum tumor marker levels. Because of the short-turnaround time and low-cost, ctDNA analysis by dPCR enables frequent and longitudinal tumor burden monitoring with a frequency similar to that for conventional serum tumor markers. Clinical validities of ctDNA and tumor markers for relapse prediction, treatment efficacy, and relapse-free corroboration were evaluated during the treatment course.

### Statistical Analysis

Through pre- and post-treatment observation, the primary goal of the present study is to assess whether ctDNA monitoring has clinical validity in terms of: prediction of early relapse, evaluation of treatment efficacy, and corroboration of relapse-free state. The ctDNA level was plotted according to VAF as a function of time. For ordinal group comparisons, Jonckheere-Terpstra test and Chi-square test were used. Correlation between two variables was calculated based on Spearman’s rank correlation coefficient. Kaplan-Meier estimation with log-rank test was used to compare OS stratified based on ctDNA level. Risk was estimated based on OS using Cox proportional hazards model. To take into account the delta value in pre- and post-treatment ctDNA levels, a landmark analysis was used for those who received treatments [17, 18]. The landmark–an arbitrary point in time after treatment–for the delta calculation was defined as the first day on which blood was drawn within 90 days of the start of first line treatment. The delta values were defined as:

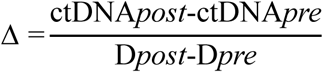

The delta level was divided into 2 categories using a delta value of 0.08% for stratified OS group comparison. Log-rank test *P* values <0.05 were considered statistically significant. All analyses were performed using GraphPad Prism 6 (GraphPad), JMP 10 (SAS) and EZR [19].

## Results

### Mutations in primary ESCC tumors

Thirty-six ESCC patients were enrolled for this study (supplementary Figure S1). One sample could not be analyzed by NGS (EC_27). Mutation screening of primary tumors in 35 patients for whom adequate tumor DNA samples were available was performed using the SCC panel. A total of 221 mutations (6.3 mutations per sample [28.4/Mb] on average) were identified that had VAF >2% (supplementary Table S3). The most frequently mutated gene was *TP53* (34/35, 97.1%; total of 45 mutations). The VAF of most other mutations was <10% (supplementary Figure S4). The patient characteristics and mutation profile are summarized in Figure 1. Sequence data were deposited in the DNA Data Bank Japan (Accession number JGAS00000000219) [20].

**Figure 1.**
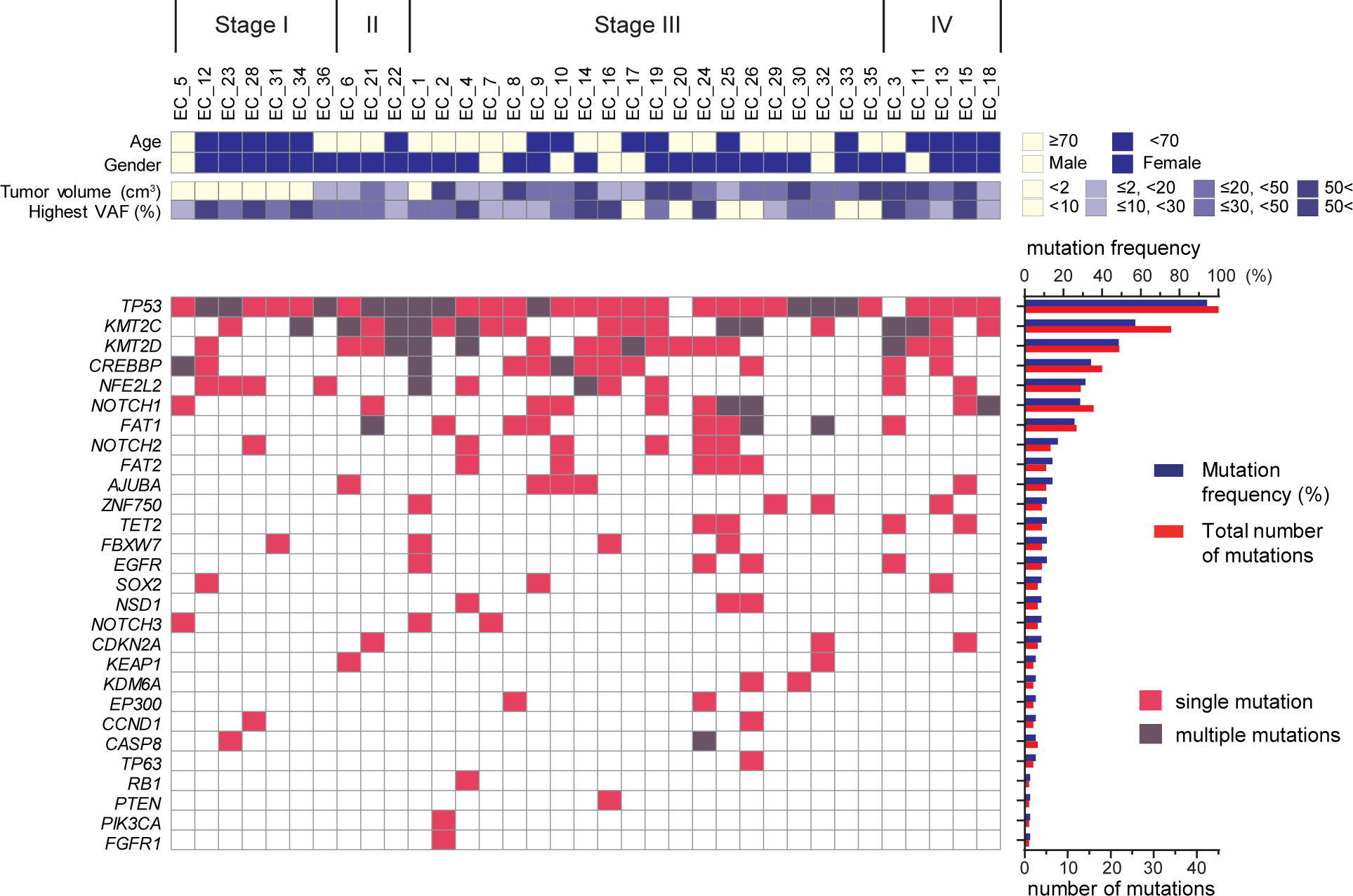
Somatic mutation profile of 35 primary ESCC tumors. Clinical characteristics are shown in the top panel. Mutated genes (>2% VAF) are shown in the bottom panel. The frequency and number of mutated genes is shown in the right side of the bottom panel. VAF, variant allele frequency.**A**

### ctDNA detection in pretreatment plasma

No probe/primer for tumor-specific mutations could be validated in tumor DNA (EC_20). Therefore, probe/primer sets for 58 selected mutations from 34 patients were validated by dPCR using primary tumor DNA and ctDNA detection at any time point (supplementary Figure S5). Strong correlations of VAFs with NGS and dPCR results were observed in primary tumors and pre-treatment plasma DNA (*r* = 0.98 and *r* = 0.72, respectively, Spearman’s rank correlation coefficient; supplementary Figure S6A, B)

In pre-treatment plasma from 34 patients, patients with advanced stage disease had higher VAFs than did early stage patients (*P* <.0001, Jonckheere-Terpstra test) (Figure 2A). The total tumor volume also correlated well with pre-treatment ctDNA VAF (*r* = 0.71, Spearman’s rank correlation coefficient, Figure 2B). Overall, the pre-treatment ctDNA-positive rate of stage I patients was 14.3% (1/7), whereas that for stage II or higher was 85.2% (23/27).

**Figure 2.**
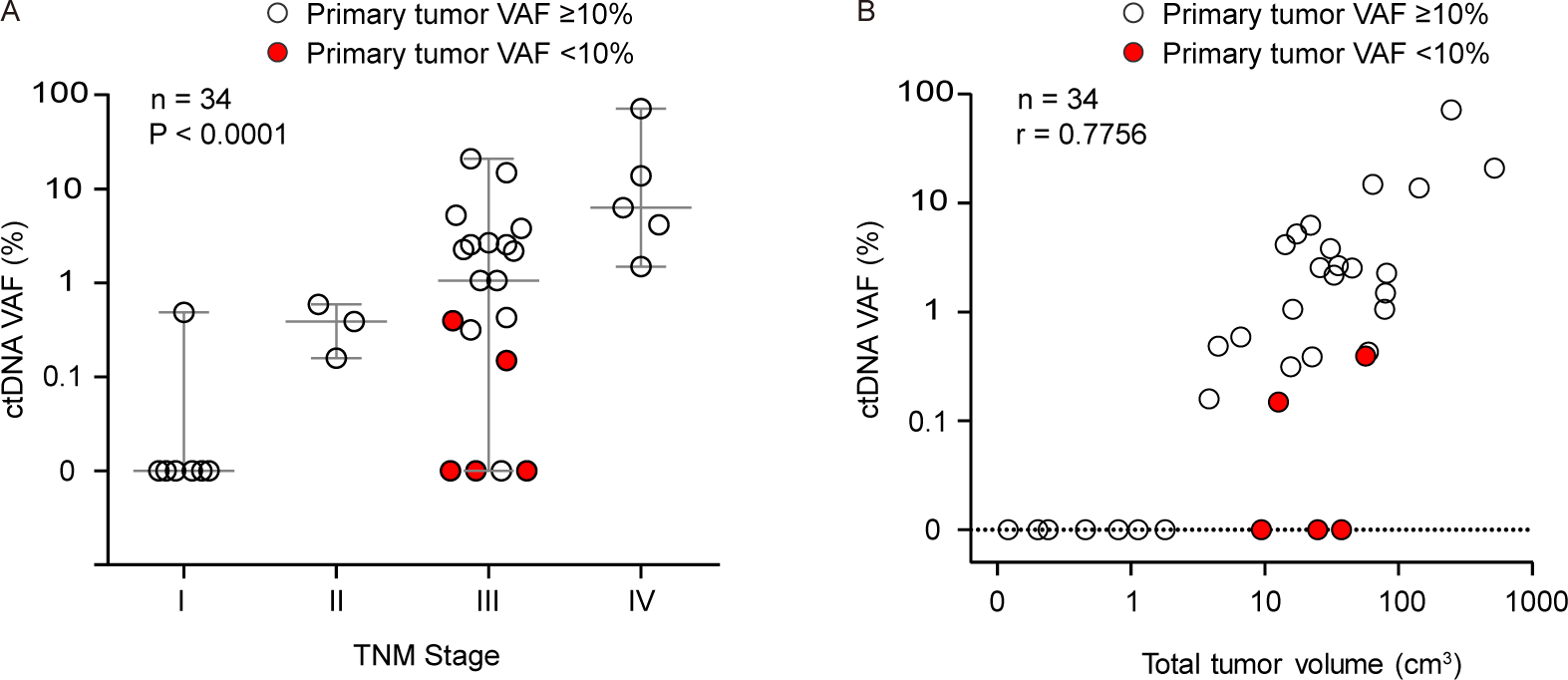
VAF of ctDNA in pre-treatment plasma in terms of TNM stage and tumor volume. (A) VAF of ctDNA in pre-treatment plasma in terms of TNM stage (P < .0001, Jonckheere-Terpstra test); (B) VAF of ctDNA in pre-treatment plasma in terms of tumor volume (r = 0.71, CI, 0.48-0.85, P < 0.0001, Spearman’s rank correlation coefficient). Open and red circles indicate mutations having primary tumor VAF of more and less than 10%, respectively. VAF, variant allele frequency; ctDNA, circulating tumor DNA.

### Early relapse prediction

In terms of the clinical validity of ctDNA as a tumor marker, EC_6 and EC_1 represent examples of early relapse prediction (Figure 3A, B) [11]. In EC_6, para-tracheal lymph node metastasis was noted by CT scan on Day 615, after 2 cycles of cisplatin/5-FU (CF) followed by surgical resection (TP 9). Notably, the level of *TP53* c.659A>G ctDNA was already elevated by Day 436, for a lead time of 179 days to imaging diagnosis (TP 7). CRT for mediastinal lymph node metastasis achieved a CR at Day 713 concomitant with the observation that ctDNA levels of both mutations had decreased below the detection limit (TP 10-13). Subsequent S-1 therapy temporarily increased levels of *TP53* c.659A>G ctDNA, likely due to the increase in cancer cell turnover, although FDG intake in the superior internal jugular region visualized by PET did not provide identification of the malignant lesion. The dynamics of *AJUBA* c.331_332insGC ctDNA appeared to reflect intra-tumor genetic heterogeneity.

**Figure 3.**
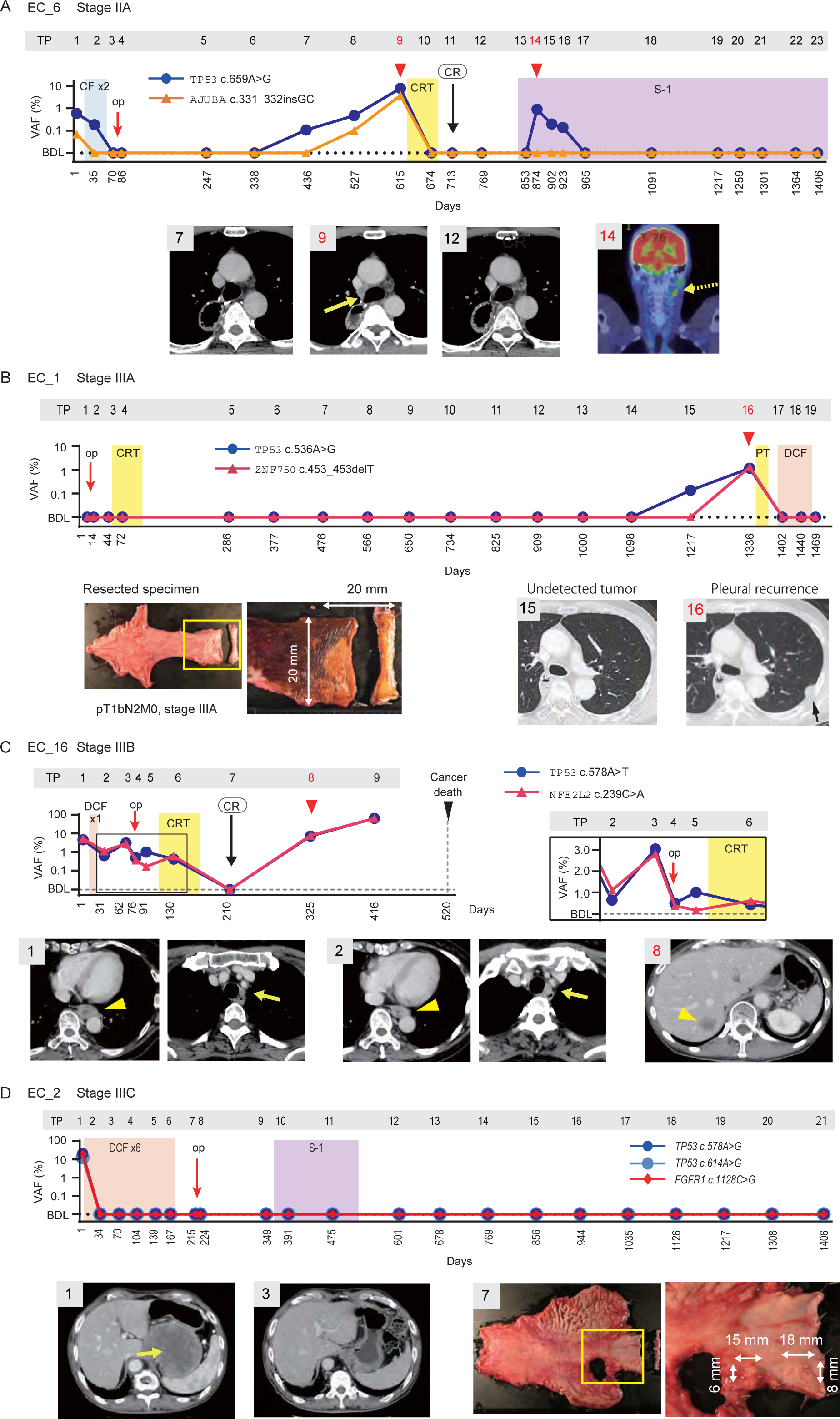
Dynamics of ctDNA during treatment course of ESCC. TP numbers on the graph indicate blood collection time points. Red arrowheads indicate clinical relapse time points. (A) EC_6, a patient having mediastinal and cervical lymph node recurrence after surgical resection. (B) EC_1 had pleural recurrence after surgical resection. (C) EC_16 showed weak response to chemotherapy. (D) EC_2 responded to chemotherapy. BDL, below detection limit; CF, cisplatin/5-FU; CRT, chemoradiotherapy; DCF, docetaxel/cisplatin/5-FU; PT, proton beam therapy; PTX, paclitaxel. TP, time point; VAF, variant allele frequency.

EC_1 did not show positive pre-treatment levels of ctDNA, while the primary tumor sequence selected 2 mutations had a sufficient level of VAFs (i.e., 24.6% for *TP53* c.536A>G; and 30.1% *ZNF750* c.453_453delT, supplementary Table S3). More than 1,000 days after surgery, the ctDNA levels in this patient remained undetectable. The *TP53* c.536A>G ctDNA was increased on Day 1,217 followed by an increase in *ZNF750* c.453_453delT ctDNA on Day 1,336 when a follow-up CT showed a nodule in the left parietal pleura. Subsequent proton beam therapy reduced the lesion volume whereupon docetaxel/cisplatin/5-FU (DCF) was given. This patient exhibited no signs of recurrence through Day 1,469. This case illustrates the importance of long-term follow-up using individual specific markers, particularly for high-risk groups. An additional 4 cases were categorized in "early relapse/progression prediction" for clinical validity (Figure 4).

**Figure 4.**
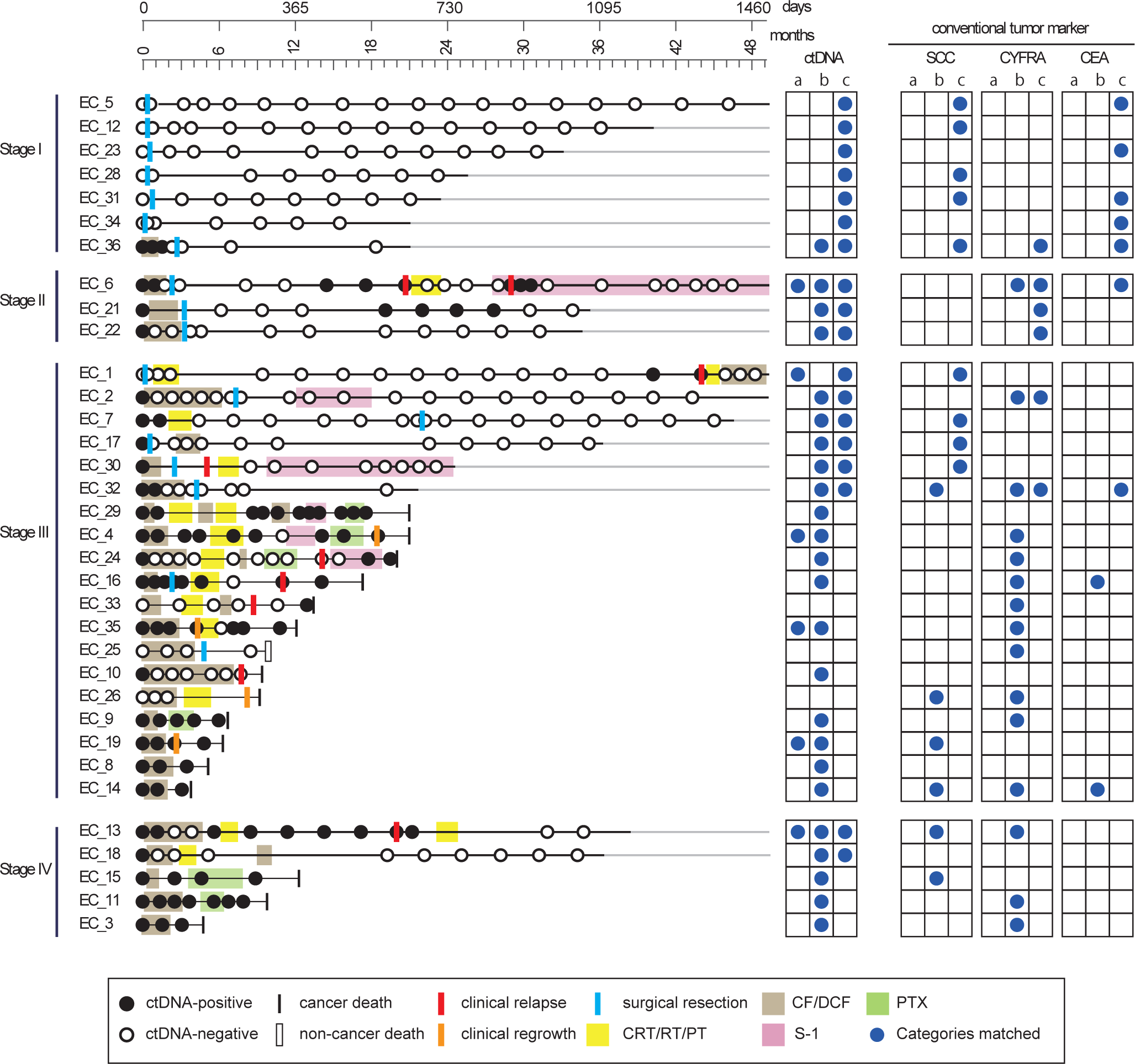
Longitudinal ctDNA monitoring and individual clinical validities. The status of ctDNA and clinical information are schematized on the horizontal lines. Blue circles in the right grid indicate representative clinical validities of ctDNA monitoring and conventional tumor markers. (a) early relapse/progression prediction; (b) treatment efficacy; (c) relapse-free corroboration. CF/DCF, chemotherapy with cisplatin/5-FU or docetaxel/cisplatin/5-FU; CRT/RT/PT, chemoradiotherapy/radiation therapy/proton beam therapy; PTX, paclitaxel.

### Treatment efficacy evaluation

The change in the level of *TP53* c.578A>T and *NFE2L2* c.239C>A ctDNA in response to chemotherapy/CRT is demonstrated by EC_16 (Figure 3C). After 1 cycle of DCF, the left recurrent nerve lymph node metastasis showed growth, although a minor response was observed in the primary tumor (TP 2). Subsequent surgical resection of the primary tumor with lymphadenectomy decreased levels of both ctDNAs. In fact, left tracheobronchial lymph node swelling was noted concurrent with *TP53* c.578A>T elevation while *NFE2L2* c.239C>A decreased, suggesting intra-tumor genetic heterogeneity (TP 5). A postoperative CRT for mediastinal metastasis resulted in CR as well as a decrease in levels of both ctDNAs to below the detection limit (TP 7). At Day 325, liver metastasis was observed by CT concomitant with >100-fold elevation in both ctDNA levels (TP 8), which both continued to increase until Day 416. At Day 520, the patient died of cancer progression. These observations suggest that both ctDNA levels can reflect treatment efficacy in a timely manner in addition to profound clonal heterogeneity in terms of treatment response. Additional 23 cases were categorized in "treatment efficacy evaluation" for clinical validity of ctDNA (Figure 4).

### Relapse-free corroboration in post-treatment follow-up

EC_2 had a primary tumor with adventitia invasion and a large abdominal lymph node metastasis that had made "retrograde" invasion to the stomach (Figure 3D). In response to one cycle of pre-operative DCF, levels of ctDNAs for *TP53* c.578A>G, *TP53* c.614A>G, and *FGFR1* c.1128C>G all dropped below the detection limit. Pathological examination of the resected specimen revealed that remnant tumor cells were limited to the submucosal layer at the gastroesophageal junction and the lower esophagus. No recurrence was observed until Day 1,406. Eighteen cases were ultimately categorized as "relapse-free corroboration" (Figure 4). Importantly, all stages were included in this category.

### Clinical validity of ctDNA in post-treatment follow-up

Our longitudinal ctDNA monitoring study was designed to confirm clinical validities [11], such as relapse/progression prediction, treatment efficacy, and relapse-free corroboration. In this study cohort, the median observation period was 638 days (range, 104 – 1,497 days). The total number of plasma samples was 569, for which 332 time points were analyzed for ctDNA. To categorize longitudinal data, the ctDNA level for each time point was binarized (i.e., ctDNA-positive and -negative) for the swimmer plot (Figure 4). In general, patients with earlier disease stages showed more frequent ctDNA-negative time points than those who had more advanced disease. Among 34 cases for which ctDNA was monitored in a longitudinal manner, 6 cases (17.6%) would have benefited from earlier relapse/progression prediction with a median lead time of 112 days over that for conventional imaging diagnosis. Meanwhile, 24 cases (70.1%) showed evaluable ctDNA dynamics in response to treatment, whereas 18 cases (52.9%) showed ctDNA-negative status over an extended period of time, confirming post-treatment relapse-free status. These clinical validations are not necessarily mutually exclusive since ctDNA may act as tumor marker at different treatment stages. Clinical validities of ctDNA were more frequently observed in longitudinal monitoring than conventional tumor markers. Of note, despite a high tumor volume, the undetectable pretreatment levels of ctDNA in three patients may have been due to low VAFs in the primary tumor (EC_25, EC_26, EC_33), and thus would not have benefited from these clinical validities. Nonetheless, the clinical validities of ctDNA monitoring were confirmed for 91% (31/34) of ESCC patients in this study. The ctDNA dynamics and detailed clinical information for all 34 patients are shown in supplementary Figure S8 (Case presentations) and Table S4.

### Overall survival by ctDNA dynamics in response to therapy

Comparison of stratified OS in terms of pre-treatment plasma showed no significant difference between patients who were ctDNA-positive and -negative (Figure 5A). This observation may be because treatment efficacy was the more predominant factor for OS than pre-treatment tumor burden. Therefore, investigation of whether patient stratification by ctDNA in terms of OS prediction by treatment response is important. A landmark approach using a pre-specified "remnant cancer landmark" was previously used to minimize guarantee-time bias [17, 18]. The "remnant cancer landmark" was defined as the blood draw taken within 3 months of the start of first line treatment. Most patients completed two cycles of chemotherapy and generally received the first follow-up scan after surgical resection by the landmark. In the present study cohort, 21 pre-treatment ctDNA-positive patients were included in the landmark analysis (supplementary Figure S1). Patients who were ctDNA-negative at the landmark (n = 10) showed significantly better OS than those who were ctDNA-positive (n = 11; *P*<.0001, HR 0.10, 95% CI, 0.03-0.30; Figure 5B). The univariate analysis showed that a decrease in ctDNA at the landmark was potentially one of the most critical prognostic factors for survival among T, N factors and pretreatment tumor volume (Supplement Table S5).

**Figure 5.**
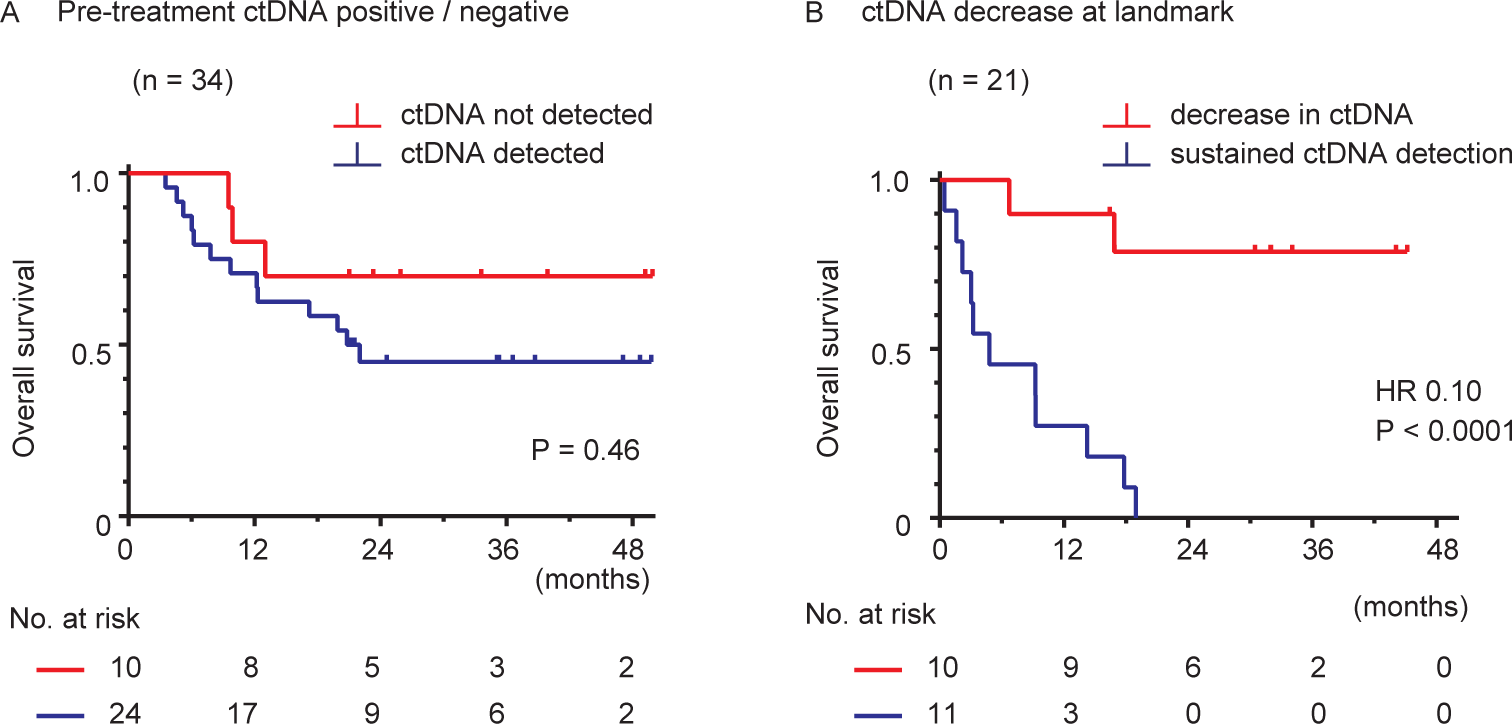
Overall survival stratified by ctDNA status. (A) OS of 34 patients stratified in terms of pretreatment ctDNA status; (B) OS for 21 patients in terms of ctDNA level at the "remnant cancer landmark". ctDNA, circulating tumor DNA; HR, hazard ratio; P values were derived from Kaplan-Meier log-rank test. HR was calculated using the log-rank test.

### *NFE2L2* mutations and response to chemotherapy

Separate from ctDNA findings, an accumulation of the *NFE2L2* mutation in the Keap1 binding domain appeared to affect chemotherapeutic response and cancer progression (supplementary Figure S7). Inhibition of the NRF2 molecule that binds to KEAP1 homodimer by either *KEAP1* or *NFE2L2* mutation is known to be involved in malignant transformation in various cancer types and resistance to chemotherapy/radiotherapy [21–27]. It has also been reported that *NFE2L2* mutations were significantly predictive and prognostic for CRT response [28]. In the present study, 10 *NFE2L2* mutations with VAF higher than 5% were found in 9 patients (9/35, 25.7%) including 4 stage I, 4 stage III, and 1 stage IV. Nine out of the 10 mutations (90%) of *NFE2L2* were located in the coding region of the KEAP1 binding domain. Five out of 6 (83.3%) mutations found in stage III, IV patients were located at the ETLG motif of *NFE2L2* whereas all mutations found in stage I (4/4, 100%) patients were located in DLG motif. These *NFE2L2* mutations were more frequently observed in non-responders to chemotherapy than responders (6 of 13 vs 0 of 14, Chi-square test, *P =* 0.039). In fact, all 5 stage III, IV and NFE2L2-mutated patients died of cancer progression within 2 years. The hazard ratio of *NFE2L2* wild type to mutation was 0.69 for overall survival (OS) (supplement Table S5).

## Discussion

This study demonstrated the clinical validity of patient-specific ctDNA for tumor burden monitoring using dPCR in standard-of-care for ESCC patients in terms of: (a) prediction of relapse/progression; (b) evaluation of treatment efficacy; and (c) corroboration of relapse-free state. Previous reports on ctDNA monitoring in ESCC or other types of cancer using NGS suggested that ctDNA could be a good tumor marker [5–9, 29] However, the cost and labor associated with NGS-based ctDNA monitoring has limited its use. To facilitate frequent monitoring in daily practice, systems to measure tumor markers should be economical, accessible, and simple to use. In the present study, we showed that frequent ctDNA monitoring using dPCR with a small number of mutations could predict clinical relapse with a median lead time of 149 days (i.e., 5 months) relative to that for conventional imaging modalities. The use of ctDNA for early detection of cancer relapse was also reported by tumor sequencing combined with dPCR on plasma samples [8, 30, 31] Results of our present study indicated that decreasing levels of ctDNA in response to chemotherapy followed by maintenance of a ctDNA-negative state predicted a longer survival time. In contrast, sustained ctDNA-positive readings in response to an initial cycle of chemotherapy suggested that the expected therapeutic efficacy would not be achieved. Together, these results indicate that frequent tumor burden monitoring using tumor-specific ctDNA by dPCR may offer opportunities for early intervention and selection of alternate, more effective relapse treatments. In addition, the high sensitivity of dPCR is a useful validation of "relapse-free" status.

A recent phylogenetic analysis of tumor ctDNA that examined 2-20 patient-specific mutations using multiregion tumor sequencing in lung cancer showed that ctDNA enabled accurate tracing of tumor burden for ctDNA-guided therapy or relapse prediction [9]. However, the experimental pipeline required both tumor and ctDNA target sequencing as well as a separate NGS run for every time point. Thus, the costs associated with the use of NGS across the overall survey period is prohibitive and would preclude its use for immediate clinical application. With dPCR, however, the number of mutations monitored is relatively limited, and NGS is not required for ctDNA monitoring [32, 33]. In this study, we first identified mutations that should be monitored were simply selected from the highest VAF within a single biopsy from a tumor. Subsequently, we monitored ctDNA by dPCR, which has a sensitivity that is generally >10-100-fold higher [34] than NGS while having a lower cost (~10 USD/assay) that allows more frequent testing. In fact, a recent report using phylogenetic tumor analysis suggested that single biopsies are likely to hold functionally important information for a limited number of mutations that in turn allows effective identification of those mutations that can serve as individual tumor markers [35]. In line with this report, the mutations that we identified from a single biopsy based on a high VAF were most likely to be "truncal" mutations. Although our system does not cover as wide a gene spectrum as NGS, a set of 1-3 mutations per patient was suitable for monitoring tumor burden in post-treatment ESCC patients.

In our present study, we used at most 30 ng DNA per dPCR run for ctDNA assay, since the detection target VAF range of ctDNA is as low as 0.01%, such that 30 ng (i.e, 10,000 human genome copy number) would be sufficient to yield a stable assay output. The amount of DNA required for that level of sensitivity can be covered from a 5 mL plasma sample isolated from 10 mL whole blood. Our post-treatment survey required only one blood collection tube per time point and this amount of blood yielded sufficient DNA for multiple replicate assays. Furthermore, dPCR offers rapid turnaround time of as little as one day. In contrast, construction of an NGS library generally requires sub-microgram amounts of DNA and the time for sequencing runs and analyses can be up to several weeks. The turnaround time of tumor markers should ideally be within one day and less than one week so prompt decisions about diagnostic/therapeutic modalities can be made. Given the potential for detection of patient-specific tumor markers, the minimal amounts of blood needed, rapid turnaround time, and overall cost, a dPCR-based approach to ctDNA monitoring is attractive for daily practice. Moreover, the short turnaround time and high sensitivity of dPCR analysis allowed evaluation of the clinical validity of ctDNA in monitoring posttreatment tumor burden. To further support the clinical utility of ctDNA, prospective randomized clinical trials are needed to determine whether therapeutic intervention with earlier relapse/progression prediction can contribute to prolonged survival.

There are several limitations to this study. As dPCR targets only known variants, undetected mutations in the tumor sequence caused by heterogeneity that arises during tumor progression cannot be evaluated. Clonal population changes in response to therapy also may not be detected with dPCR based on mutational data from a single biopsy. Another limitation of our system in a clinical setting is that the temporary cost and time needed for synthesis of specifically-designed probes for each tumor-specific mutation. In this study, however, 6 *TP53* and a *NFE2L2* recurrent mutations were observed among 17 patients (supplementary Table S6). Thus, off-the-shelf primer/probe sets could be constructed for dPCR that would cover a wide range of human cancers in post-therapeutic surveys.

In conclusion, here we demonstrated the clinical validity of tumor burden monitoring of individualized ctDNA using tumor sequencing in combination with plasma DNA dPCR as a standard-of-care framework for ESCC patients.

## Data Availability

The authors confirm that the data supporting the findings of this study are available within the article [and/or] its supplementary materials.

## Acknowledgements

We thank the surgeons in Department of Surgery, Iwate Medical University, Drs. S. Baba, A. Umemura, H. Nikai, K. Sato, and T. Chiba, who contributed patients.

